# Genetic Variants and Alteration in *Transcription Factor 7-Like 2 (TCF7L2)* mRNA level in Ischemic Stroke Patients: An Indian Scenario

**DOI:** 10.1101/2024.12.09.24318748

**Authors:** Dipanwita Sadhukhan, Arunima Roy, Sukla Nath, Esha Basu, Joydeep Mukherjee, Kartik Chandra Ghosh, Tapas Kumar Banerjee, Prasad Krishnan, Sourya Kishore Chatterjee, Soaham Desai, Vishal Ajitbhai Patel, Debes Ray, Subhra Prakash Hui, Soma Gupta, Arindam Biswas

**Affiliations:** Molecular Biology & Clinical Neuroscience Division, National Neurosciences Centre, Calcutta, Kolkata, India; Department of Biochemistry, Nilratan Sircar Medical College, Kolkata, India; Department of Neurology, Nilratan Sircar Medical College, Kolkata, India; Department of NeurologyNational Neurosciences Centre, Calcutta, Kolkata, India; Dept of Neurology, Shree Krishna Hospital and Pramukhswami Medical College, Bhaikaka University, Karamsad, Anand, Gujarat, India; S. N. Pradhan Centre for Neurosciences, University of Calcutta, Kolkata, India; DBT-NIDAN Kendra, Nilratan Sircar Medical College, Kolkata, India

**Author notes:** Corresponding author: Dr. Arindam Biswas Molecular Biology & Clinical Neuroscience Division National Neurosciences Centre, Calcutta Peerless Hospital (2n^d^ floor) 360, Panchasayar, Kolkata 700094 India Dr. Dipanwita Sadhukhan Molecular Biology & Clinical Neuroscience Division National Neurosciences Centre, Calcutta Peerless Hospital (2^nd^ floor) 360, Panchasayar, Kolkata 700094 India.

**Keywords:** *TCF7L2*, Transcript analysis, Genetic Variants, Ischemic Stroke, Hyperglycemia, Hyperlipidaemia, Indians

## Abstract

**Background:** Diabetes and Hyperlipidemia are major risk factors for stroke across the world population. *TCF7L2*, a key player of the WNT signaling pathway shows genetic association with both metabolic disturbances. However, its role in stroke pathogenesis (if any) is not well characterized.

**Objectives:** Thus, here we aim to (a) examine and correlate dysregulation of *TCF7L2* expression with diabetes or hyperlipidemia-associated Ischemic Stroke, (b) identify genetic risk variants in the *TCF7L2* gene and (c) establish a correlation between *TCF7L2* mRNA expressions with biochemical parameters.

**Methods:** Based on radiological findings for Ischemic Stroke, a total of 50 unrelated subjects were recruited with diverse biochemical parameters for TCF7L2 mRNA expression study in PBMC, followed by correlation with fasting blood sugar and lipid profile. Furthermore, mutation screening and genetic association studies (rs7901695 & rs7903146) were performed among 326 cases and 258 controls from India.

**Results:** Here, we observed a significant downregulation of *TCF7L2* gene expression among hyperlipidemic stroke patients than cases and control without dyslipidemia but no change between the cases with different diabetic statuses. Moreover, a strong negative correlation between *TCF7L2* mRNA level and total blood cholesterol but not for FBS was identified. The rs7901695T/C appeared as a promising genetic risk factor for stroke among eastern Indians.

**Conclusion:** Therefore, we can suggest that alteration in *TCF7L2* leading to stroke pathogenesis is more associated with hyperlipidemia than diabetes among Indians. Its expression level in PBMC is influenced by rs7901695T/C and holds a good promise to be used as a diagnostic marker.

## Introduction

Stroke remains a critical global health challenge, representing a leading cause of mortality and long-term disability worldwide. Metabolic risk factors, particularly hyperglycemia and hyperlipidemia, play a pivotal role in stroke pathogenesis, demonstrably increasing stroke risk by 1.5-2 times through complex pathological alterations in cerebrovascular structures (1, 2).

Recent genetic epidemiological research has illuminated the intricate relationship between genetic predisposition and stroke risk. Specifically, genetic susceptibility to Type 2 Diabetes (T2D) has been consistently associated with elevated stroke risk, particularly among populations of European ancestry (3). This genetic complexity underscores the need for comprehensive investigations across diverse ethnic populations.

*Transcription factor 7-like 2* (*TCF7L2*), localized on chromosome 10q25.3, emerges as a critical molecular mediator in this context. As a key component of the Wnt signaling pathway, *TCF7L2* plays a crucial role in regulating pro-insulin production, processing, and adipogenesis (4). Multiple polymorphic variants of *TCF7L2*, particularly rs7903146 and rs12255372, have demonstrated significant associations with Type 2 Diabetes Mellitus (T2DM) and metabolic dysregulation across various ethnic groups, including Indian populations (5–8).

Despite growing evidence linking *TCF7L2* to metabolic disorders, the molecular mechanisms underlying its potential contribution to stroke pathogenesis remain incompletely understood. Notably, only a single study has explored the genetic association of *TCF7L2* with diabetes-induced stroke, and this was limited to a Korean population (5), highlighting a substantial research gap.

Given the background, we hypothesize that *dysregulation of TCF7L2 expression and its genetic variants may contribute to stroke susceptibility by modulating glucose and lipid metabolic pathways*.

Addressing these critical knowledge gaps, our study aims to (a) Comprehensively examine *TCF7L2* transcript level alterations in stroke patients, stratified by diabetic and lipid metabolic profiles, (b) Conduct genetic association studies with functional *TCF7L2* variants and (c) Investigate the potential impact of metabolic parameters on ischemic stroke lesion characteristics and subtypes within an Indian cohort

## MATERIALS AND METHODS

### Study Population and Design

This multicentre, case-control study was conducted across three prominent medical institutions in India: National Neurosciences Centre, Calcutta; Nil Ratan Sircar Medical College & Hospital, Kolkata, and Shree Krishna Hospital, Pramukhswami Medical College, Gujarat. The study cohort comprised 326 ischemic stroke patients (252 from Kolkata and 74 from Gujarat) diagnosed with various stroke subtypes, including small vessel disease, large vessel disease, and cardio-embolism, confirmed through comprehensive radiological imaging. Participants were carefully selected based on stringent inclusion and exclusion criteria to ensure methodological rigor and scientific validity.

The inclusion criteria encompassed patients with radiologically confirmed ischemic stroke, diabetes diagnosed by HbA1c levels prior to stroke onset, age range of 30-85 years, and diabetes duration less than 5 years. Exclusion criteria eliminated potential confounding factors by excluding individuals with a cancer history, type 1 or gestational diabetes, and pregnant or lactating women. A control group of 258 neurologically healthy individuals (mean age 57.83 ± 10.55 years) without personal or familial stroke history was recruited from Kolkata to facilitate comparative analysis. The demographic details of the subjects are described in Table 1A. Each patient was assessed in detail by history from a reliable caregiver and neurological examination for primary assessment.

**Table 1A:**
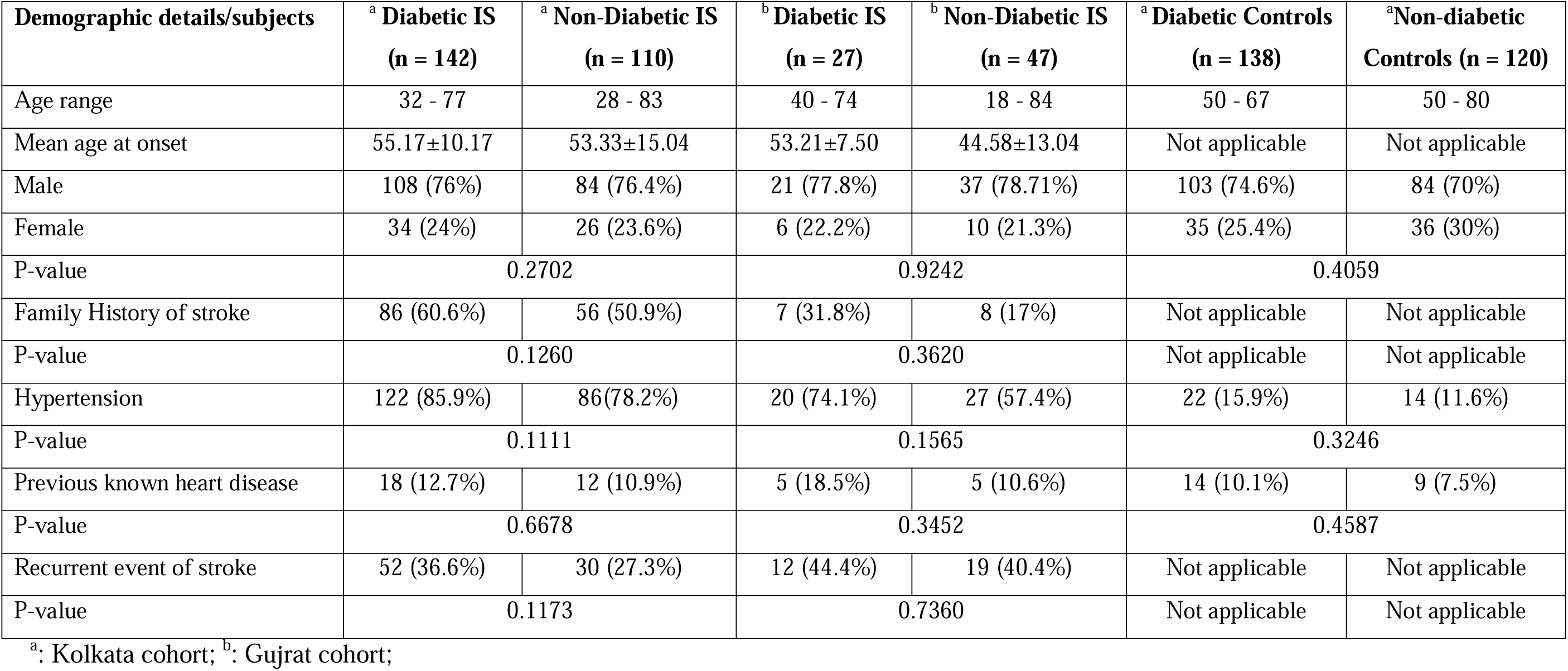
Demographic details of study subjects.

| **Demographic details/subjects** | **<sup>a</sup> Diabetic IS**<br>**(n = 142)** | **<sup>a</sup> Non-Diabetic IS**<br>**(n = 110)** | **<sup>b</sup> Diabetic IS**<br>**(n = 27)** | **<sup>b</sup> Non-Diabetic IS**<br>**(n = 47)** | **<sup>a</sup> Diabetic Controls**<br>**(n = 138)** | **<sup>a</sup>Non-diabetic**<br>**Controls (n = 120)** |
| --- | --- | --- | --- | --- | --- | --- |
| Age range | 32 - 77 | 28 - 83 | 40 - 74 | 18 - 84 | 50 - 67 | 50 - 80 |
| Mean age at onset | 55.17±10.17 | 53.33±15.04 | 53.21±7.50 | 44.58±13.04 | Not applicable | Not applicable |
| Male | 108 (76%) | 84 (76.4%) | 21 (77.8%) | 37 (78.71%) | 103 (74.6%) | 84 (70%) |
| Female | 34 (24%) | 26 (23.6%) | 6 (22.2%) | 10 (21.3%) | 35 (25.4%) | 36 (30%) |
| P-value | 0.2702 |  | 0.9242 |  | 0.4059 |  |
| Family History of stroke | 86 (60.6%) | 56 (50.9%) | 7 (31.8%) | 8 (17%) | Not applicable | Not applicable |
| P-value | 0.1260 |  | 0.3620 |  | Not applicable | Not applicable |
| Hypertension | 122 (85.9%) | 86(78.2%) | 20 (74.1%) | 27 (57.4%) | 22 (15.9%) | 14 (11.6%) |
| P-value | 0.1111 |  | 0.1565 |  | 0.3246 |  |
| Previous known heart disease | 18 (12.7%) | 12 (10.9%) | 5 (18.5%) | 5 (10.6%) | 14 (10.1%) | 9 (7.5%) |
| P-value | 0.6678 |  | 0.3452 |  | 0.4587 |  |
| Recurrent event of stroke | 52 (36.6%) | 30 (27.3%) | 12 (44.4%) | 19 (40.4%) | Not applicable | Not applicable |
| P-value | 0.1173 |  | 0.7360 |  | Not applicable | Not applicable |

<sup>a</sup>: Kolkata cohort; <sup>b</sup>: Gujrat cohort;

### Determination of Biochemical parameters

Biochemical parameters were comprehensively analyzed using the Roche Cobas c-501 Automatic Analyzer, quantifying crucial metabolic markers including fasting plasma glucose, glycated hemoglobin, serum total cholesterol, triglycerides, and lipid profile (HDL and LDL) (9).

### Gene Expression Analysis

#### Sample collection

For molecular analysis, peripheral blood mononuclear cells (PBMCs) were isolated from 50 participants using Histopaque 1077 double-gradient density centrifugation, with blood samples collected from fasting individuals between 10:30 AM and 11:30 AM to minimize diurnal variations.

#### RNA extraction, cDNA synthesis, and quantitative PCR

Total RNA extraction was performed using Trizol reagent (Thermo Fisher Scientific, US), with 2 µg of RNA subjected to cDNA synthesis using a reverse transcriptase kit (Promega Corporation, USA) following standard procedures. Quantitative real-time PCR (qPCR) was conducted to quantify *TCF7L2* gene expression using specific primers (forward primer: 5′-CATATGGTCCCACCACATCA-3′, reverse primer 5′-CACTCTGGGACGATTCCTGT-3′), with 18S rRNA serving as an endogenous control (Forward primer: 5’-GTAACCCGTTGAACCCCATT-3’, reverse primer 5’-CCATCCAATCGGTAGTAGCG-3’). The QuantStudio-5 Real-Time PCR system (Applied Biosystems, US) facilitated precise quantification of gene expression levels.

### Genetic Variant Analysis

#### *TCF7L2* mutation screening

Genomic DNA was extracted using a QIAamp DNA Blood kit (QIAGEN, Germany). All the coding regions of the *TCF7L2* gene were sequenced by targeted gene capture using a custom capture kit. Paired-end sequencing was performed with 2X100/2X150 chemistry in the Illumina NovaSeq 6000 sequencing platform. Reads were assembled and aligned to reference sequence based on NCBI RefSeq transcript and human genome build GRCh37/UCSC hg19.

#### Genetic Association study

Based on an extensive literature review, two single nucleotide polymorphisms (SNPs) were selected for detailed investigation: rs7901695 T/C and rs7903146 C/T, previously associated with type 2 diabetes, dyslipidemia, and diabetes-induced stroke in diverse populations (4).

Genomic DNA was isolated from 5 mL peripheral blood samples, with target regions amplified through PCR and subsequently analyzed using restriction fragment length polymorphism (RFLP) techniques. The RFLP analysis employed BstUI and HpyCHIV-III restriction enzymes (New England Biolabs, USA), with variant identification confirmed through bidirectional Sanger Sequencing (∼20% of RFLP data). This multi-step approach ensured robust and reliable genetic variant detection.

### Statistical Analysis

Statistical analysis was performed using multiple computational approaches to ensure comprehensive and rigorous data interpretation. Demographic and clinical parameters were evaluated using Mann-Whitney U test and Fisher exact test, with statistical significance set at p ≤ 0.05. Genotype associations were examined using odds ratios and 95% confidence intervals, with the JavaStat online tool facilitating complex statistical calculations (http://statp ages. info/ ctab2 x2. html).

To mitigate multiple testing challenges, the Benjamini-Hochberg correction method was applied to control the false discovery rate. Gene expression data, correlation analyses, and receiver operating characteristic (ROC) curve analyses were conducted using GraphPad Prism 5.0. Data were presented as mean log_2_-transformed expression ± Standard Error of Mean (SEM), providing a comprehensive statistical framework for interpreting genetic and molecular findings.

### Ethical Considerations

The entire research protocol was conducted in strict accordance with the guidelines of the Indian Council of Medical Research (ICMR). Informed consent was obtained from all participants, ensuring ethical standards were meticulously maintained throughout the study.

## RESULTS

### Demographic Characteristics of Study Subjects

The study recruited a total of 326 ischemic stroke cases (252 from Kolkata and 74 from Gujrat) and 258 age- and ethnicity-matched controls (Kolkata) from India. The mean age of onset for cases was 51.17±10.17 years, with a male-to-female ratio of 3.29:1. The control group had a mean age of 57.42±9.35 years and a male-to-female ratio of 2.63:1 (Table 1A).

### Biochemical Parameters in Ischemic Stroke Subjects

Compared to controls, diabetic stroke cases had significantly higher fasting blood sugar, total cholesterol, and LDL-cholesterol levels (P values = 0.0005407, 0.00553, and 0.01041, respectively). In non-diabetic subjects, total cholesterol and LDL-cholesterol levels were also elevated compared to controls (P values = 0.002689 and 0.004804, respectively), but fasting blood sugar was not significantly different (Table 1B).

**Table 1B:** Comparison of biochemical parameters among the study cohort.

|  | **Fasting blood sugar** | **Total Cholesterol** | **Triglyceride** | **HDL** | **LDL** |
| --- | --- | --- | --- | --- | --- |
| **Diabetic Ischemic Stroke** | 184.7±73.54 | 168.57±50.56 | 138.41±47.39 | 43.44±8.41 | 101±39.95 |
| **Diabetic controls** | 137.04±34.91 | 145.57±29.94 | 155.90±59.22 | 47.95±11.79 | 67.18±24.93 |
| **P-Value** | **0.0005407** | **0.00553** | 0.1907 | 0.2499 | **0.01041** |
| **Non-Diabetic Ischemic Stroke** | 98.12±9.45 | 173.83±46.83 | 141.32±47.29 | 55.2±19.65 | 122±30.98 |
| **Non-Diabetic Controls** | 97.21±20.95 | 137.85±27.62 | 140.27±58.09 | 44.43±9.52 | 68.45±27.69 |
| **P-Value** | 0.9085 | **0.002689** | 0.7115 | 0.2972 | **0.004804** |

### Association of Biochemical Parameters with Arterial Territory Involvement

Ischemic stroke cases were stratified based on the major arterial territory involved (middle cerebral artery infarcts, n = 184; posterior cerebral artery infarcts, n = 57). However, no significant associations were found between elevated fasting blood sugar or lipid levels and specific arterial territory involvement (data not shown).

### *TCF7L2* Expression in Ischemic Stroke

Gene expression analysis of *TCF7L2* was performed in 50 study subjects (29 cases, 21 controls). Irrespective of diabetic status, *TCF7L2* expression was significantly downregulated in ischemic stroke cases compared to controls (Figure 1A-C). However, no significant difference in *TCF7L2* expression was observed between diabetic and non-diabetic stroke cases (Figure 1C). It is clear from Figure 1A and Figure 1B, that irrespective of diabetic status among stroke cases, *TCF7L2* expression was found to be downregulated than respective control groups. For diabetic subjects, mean log2 *TCF7L2* expression in cases and controls were = −2.782± 0.08624, −2.205± 0.1299 respectively; P = 0.0027 (Figure 1A) while for non-diabetic subjects the values were − 3.078 ± 0.1154, −2.376 ± 0.07139, P = 0.0001 (Figure 1B). However, no significant difference was observed between diabetic Ischemic Stroke and non-diabetic Ischemic Stroke cases (P = 0.2248; Figure 1C).

**Figure 1:**
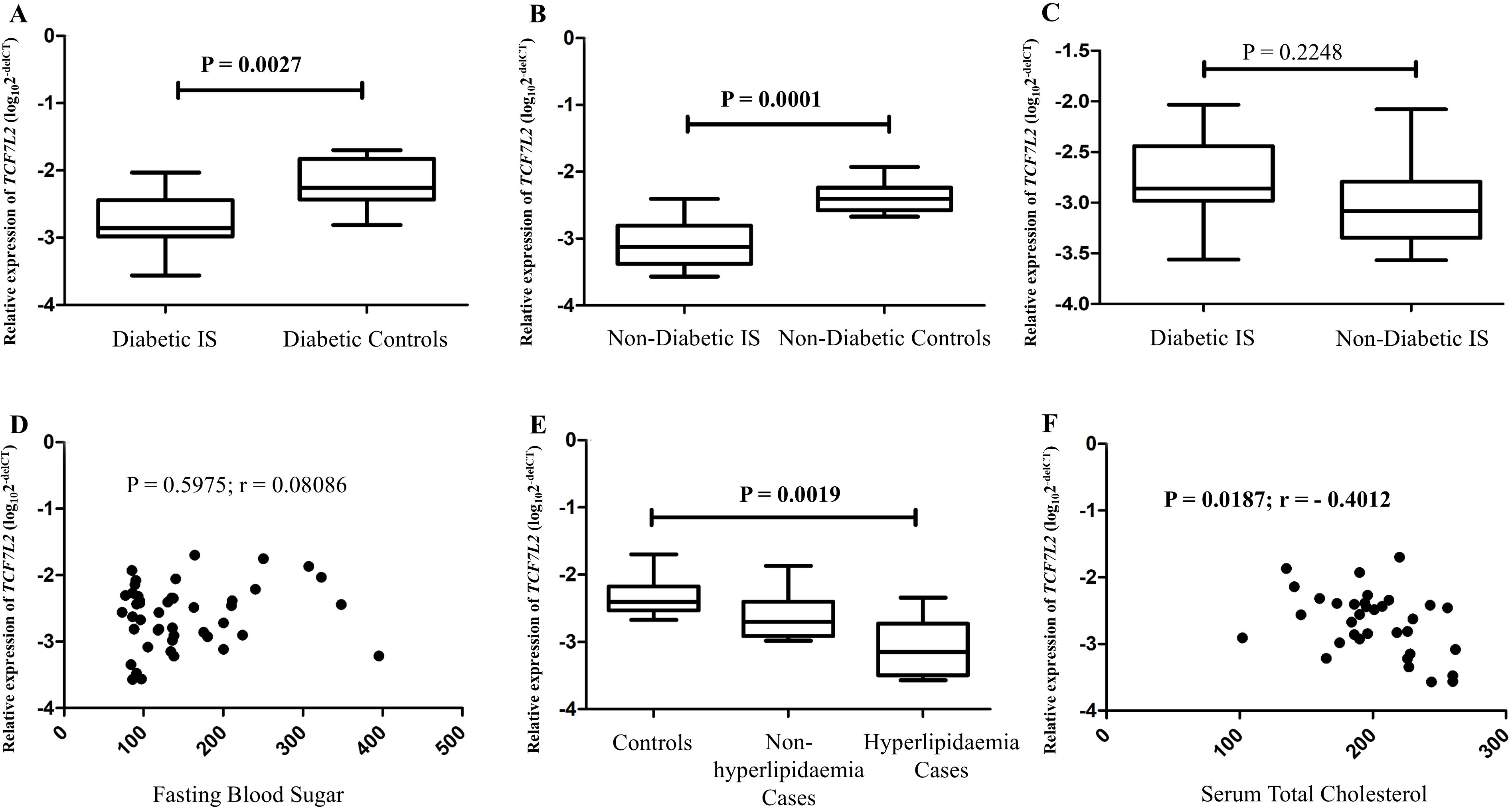
The alteration of *TCF7L2* gene expression among Ischemic Stroke patients and its correlation with fasting blood sugar and plasma total cholesterol levels. **A, B, C:** Quantification of *TCF7L2* genes in PBMC **A)** Diabetes Ischemic Stroke patients (n = 19) and Diabetes Controls (n = 11); **B)** Non-diabetes Ischemic Stroke (n = 10) and Non-diabetic Controls (n = 10); **C)** Diabetic Ischemic Stroke (n = 19) and Non-Diabetic Ischemic Stroke (n = 10) respectively. **D)** Correlation of *TCF7L2* gene expression with FBS values (n = 43). **E)** Quantification of *TCF7L2* mRNA in PBMC among controls (n = 12), Ischemic Stroke with hyperlipidaemia (n = 13) and Ischemic Stroke with without hyperlipidaemia (n = 13). **F)** Significant negative Correlation of *TCF7L2* gene expression with total cholesterol values (n = 34). *TCF7L2* gene expression level in all cases were normalised to 18S. The comparison was done by using Mann-Whitney U test and Kruskal-Wallis test; the significance level was set at P ≤ 0.05.

### Correlation of *TCF7L2* mRNA Expression with FBS

There was no significant correlation between *TCF7L2* expression and fasting blood sugar levels (P = 0.5975; r = 0.08086) [Figure 1D].

### Correlation of *TCF7L2* mRNA Expression with Total Cholesterol Levels

It is clear from Table 1B, that the mean values for total cholesterol levels are almost similar (168.57±50.56 vs 173.83±46.83; P = 0.7785) between diabetic and non-diabetic Ischemic Stroke cases. Thus, the diabetic status was not considered when cases were stratified based on blood lipid level followed by the comparative analysis.

A statistically significant downregulation in *TCF7L2* mRNA expression was observed in cases with hyperlipidaemia than in control and cases without dyslipidemia. The mean log_2_ *TCF7L2* expression among hyperlipidaemic (n = 13), non-dyslipidamia cases (n = 13) and control (n = 12) were -3.071 ± 0.1395, -2.618 ± 0.1114, -2.326± 0.0823 respectively; P = 0.0019 (Figure 1E).

### *TCF7L2* mRNA Expression in Correlation to Blood Total Cholesterol Value

Next, to investigate whether *TCF7L2* gene expression can be linked to total cholesterol level among stroke cases (n = 34), a Spearman analysis was performed. Here we found that unlike FBS the *TCF7L2* gene expression was significantly negatively correlated with the total cholesterol (r = - 0.4012, 95%CI = - 0.6570 to - 0.06244, P = 0.0187) [Figure 1F].

### *TCF7L2* as a Diagnostic Marker for Ischemic Stroke

Receiver operating characteristic (ROC) curve analysis showed that *TCF7L2* mRNA level had a good predictive value for diagnosing ischemic stroke, with an area under the curve of 0.8346±0.07163 (95% CI = 0.6942 – 0.9750, P = 0.001357) (Figure 2). The optimal cut-off value for *TCF7L2* mRNA was -2.559, with a sensitivity of 0.75 and a specificity of 0.77. Among the 47 individuals recruited for mRNA expression study, 66% of subject representing cases and 20% individuals representing controls showed downregulation of *TCF7L2*.

**Figure 2:**
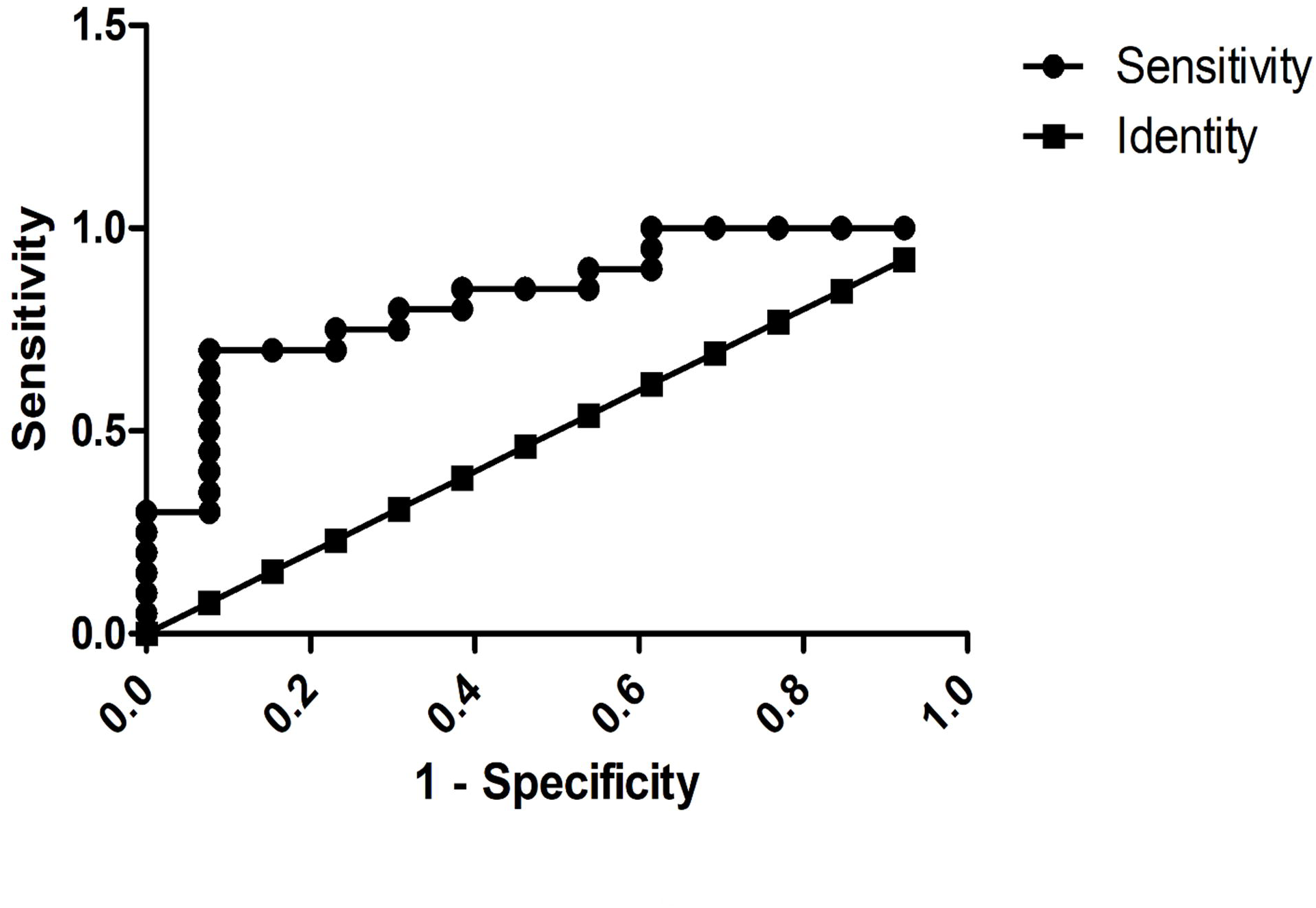
ROC curve for PBMC *TCF7L2* mRNA level as diagnostic marker for Stroke.

### Genetic Variants in *TCF7L2*

Screening of the TCF7L2 coding region in 31 ischemic stroke cases and 30 controls identified one rare missense variant (rs1234434500, c.70G>A, p.Gly24Arg) and two other variants (rs201875922, c.948A>G, p.Thr316Thr; rs77961654 in the 3’UTR region) in the stroke cases. Except for the Gly24Arg variant, the other two variants were also found in the control group at similar frequencies.

### Genetic Association of *TCF7L2* Variants with Diabetes-related Ischemic Stroke

Next, to explore the genetic association of *TCF7L2* with diabetes and hyperlipidemia associated stroke pathomechanisms, intronic yet functional variants like rs7901695 and rs7903146 were genotyped in 142 unrelated diabetics Ischemic Stroke cases, 138 diabetic controls, 110 non-diabetics Ischemic Stroke cases and 120 non-diabetics controls of same ethnicity from Eastern India. Both the SNVs were not in linkage disequilibrium (LD) either in patient group or control groups (r^2^ value = 47, 20 respectively).

For the rs7901695, C-allele [Odds ratio = 1.5298; 95% CI = 1.067-2.314; P = 0.0207] and CC+TC genotype [Odds ratio = 2.5379; 95%CI = 1.566 - 4.112; P = 0.0002] were observed to be associated with risk for only diabetes-related stroke but not for non-diabetic stroke [P = 0.6795] [Table 2]. Furthermore, a statistically significant difference was also identified for this ‘C’ allele [Odds ratio = 2.119; 95% CI = 1.413-3.179; P = 0.0003] and CC+TC genotype [Odds ratio = 2.760; 95%CI = 1.651 - 4.614; P = 0.0001] between diabetic cases and non-diabetic cases [Table 2]. Interestingly, the T-allele of rs7901695 was found to be overrepresented among stroke subjects with hyperlipidaemia than cases with low cholesterol level [Odds ratio = 1.5942; 95% CI = 1.1.0487 – 2.4235; P = 0.0291 and Odds ratio = 1.6941; 95%CI = 1.0051 – 2.8555; P = 0.0478 respectively] [Table 3].

**Table 2:** Genotype and Allele frequency distribution of *TCF7L2* gene polymorphism (rs7901695 & rs7903146) among study cohort.

| **rs7901695 (Population from West Bengal)** |  |  |  |  |  |  |  |
| --- | --- | --- | --- | --- | --- | --- | --- |
| Study Subjects / Genetic variants | Genotype frequency |  |  | Allele frequency |  | P-value | Odds ratio (95% CI) |
|  | TT | TC | CC | T | C |  |  |
| **\*Diabetic IS (n = 142)** | 50 (35.2%) | 82 (57.7%) | 10 (7.1%) | 182 (64.1%) | 102 (35.9%) | **C vs T: 0.0207/\*0.0003** | **1.5298 (1.067 – 2.314)/\*2.1199 (1.413 – 3.179)** |
| **Diabetic Controls (n = 138)** | 80 (58%) | 42 (30.4%) | 16 (11.6%) | 202 (73.2%) | 74 (26.8%) | **CC+TC vs TT: 0.0002/\*0.0001** | **2.5379 (1.566 – 4.112)/\*2.760 (1.651 – 4.614)** |
| **\*Non-diabetic IS (n = 110)** | 66 (60%) | 42 (38.2%) | 2 (1.8%) | 174 (79.1%) | 46 (20.9%) | C vs T: 0.6795 | 0.9106 (0.5840 – 1.4200) |
| **Non-diabetic Controls (n = 120)** | 78 (65%) | 30 (25%) | 12 (10%) | 186 (77.5%) | 54 (22.5%) | CC+TC vs TT: 0.4340 | 1.2381 (0.7251 – 2.1140) |
| **rs7903146 (Population from West Bengal)** |  |  |  |  |  |  |  |
|  | CC | TC | TT | C | T | P-value | Odds ratio (95% CI) |
| **Diabetic IS (n = 114)** | 57 (50%) | 48 (42.1%) | 9 (7.9%) | 162 (71.1%) | 66 (28.9%) | T vs C: 0.3764 | 0.8420 (0.5751 – 1.233) |
| **Diabetic Controls (n = 138)** | 60 (43.5%) | 66 (47.8%) | 12 (8.7%) | 186 (67.4%) | 90 (32.6%) | TT+TC vs CC: 0.3019 | 0.7692 (0.4675 – 1.266) |
| **Non-Diabetic IS (n = 60)** | 30 (50%) | 28 (46.6%) | 2 (3.4%) | 88 (73.4%) | 32 (26.6%) | T vs C: 0.4650 | 0.8286 (0.5004 – 1.3720) |
| **Non-diabetic Controls (n = 100)** | 49 (49%) | 41 (41%) | 10 (10%) | 139 (69.5%) | 61 (30.5%) | TT+TC vs CC: 0.9025 | 0.9608 (0.5065 – 1.8224) |
| **rs7901695 (Population from Gujrat)** |  |  |  |  |  |  |  |
|  | CC | TC | TT | C | T | P-value | Odds ratio (95% CI) |
| **Diabetic IS (n = 27)** | 9 (33.33%) | 16 (59.27%) | 2 (7.4%) | 34 (63%) | 20 (37%) | C vs T: 0.0666 | 1.9786 0.9544 – 4.1021) |
| **Non-Diabetic IS (n = 47)** | 29 (59.6%) | 16 (34%) | 3 (6.4%) | 72 (76.6%) | 22 (23.4%) | CC+TC vs TT: **0.0267** | **3.0526 (1.1373 – 8.1939)** |
| **rs7903146 (Population from Gujrat)** |  |  |  |  |  |  |  |
|  | CC | TC | TT | C | T | P-value | Odds ratio (95% CI) |
| **Diabetic IS (n = 27)** | 12 (44.4%) | 14 (52%) | 1 (3.6%) | 38 (70.4%) | 16 (29.6%) | T vs C: 0.0936 | 1.9567 (0.8927 – 4.2886) |
| **Non-Diabetic IS (n = 48)** | 35 (73%) | 9 (18.7%) | 4 (8.3%) | 79 (82.3%) | 17 (17.7%) | TT+TC vs CC: **0.0164** | **3.3654 (1.2496 – 9.0635)** |

\*Diabetic Ischemic Stroke vs Non-Diabetic Ischemic Stroke

**Table 3:** Genotype and Allele frequency distribution of *TCF7L2* gene polymorphism (rs7901695) based on lipid profile.

| **rs7901695 (Population from West Bengal)** |  |  |  |  |  |  |  |
| --- | --- | --- | --- | --- | --- | --- | --- |
| **Study Subjects / Genetic variants** | **Genotype frequency** |  |  | **Allele frequency** |  | **P-value** | **Odds ratio (95% CI)** |
|  | **TT** | **TC** | **CC** | **T** | **C** |  |  |
| **IS with Hyperlipidaemia (n = 88)** | 48 (54.5%) | 39 (44.3%) | 1 (1.2%) | 135 (76.7%) | 41 (23.3%) | **T vs C: 0.0291** | **1.594 (1.049 – 2.424)** |
| **IS with non-Hyperlipidaemia (n = 164)** | 68 (41.5%) | 85 (51.8%) | 11 (6.7%) | 221 (67.4%) | 107 (32.6%) | **TT vs CC+TC: 0.0478** | **1.694 (1.005 – 2.855)** |

On the other hand, for rs7903146 neither allelic nor genotype was found to be associated with the disease irrespective of diabetic status [Table 2]. Because two SNVs were examined in the present study, after doing FDR (false discovery rate) for multiple testing, the significance of allelic association between the ‘C’ allele of rs7901695 and diabetes-induced stroke withstand multiple testing (P = 0.0207, which is less than the adjusted threshold P-value of 0.025 for two SNVs).

Haplotypes, were next determined based on the genotypes at rs7901695 and rs7903146 for each individual using Haploview (version 4.2). However, neither protective nor risk haplotype was identified in our study cohort (data not shown).

Likewise, for the Ischemic Stroke cases from Gujrati population both the SNVs were found to be overrepresented among diabetic Ischemic Stroke cases than non-diabetic cases [For rs7901695: P value = 0.0267; OR = 3.0526; 95% CI: 1.1373 – 8.1939 and for rs7903146: P=0.0164; OR=3.3654, 95%CI= 1.2496 – 9.0635) [Table 2].

Next, to find the impact of the genotypes on biochemical parameters considered above, subjects were stratified accordingly. However, only significant lowering in FBS value was observed for ‘TT’ genotype of rs7901695 among diabetic controls (P = 0.03443) [Table 4].

**Table 4:** Comparison of biochemical parameters based on rs7901695 genotypes.

|  |  | **TT** | **TC+CC** | **P-value** |
| --- | --- | --- | --- | --- |
| **Diabetic Controls** | Fasting blood sugar | 128.5±24.18 | 151.52±48.15 | **0.03443** |
|  | Total Cholesterol | 144.51±32.07 | 152.03±33.09 | 0.3524 |
|  | Triglyceride | 150.92±52.91 | 174.05±81.11 | 0.3586 |
| **Diabetic IS** | Fasting blood sugar | 181.86±93.72 | 185.14±67.42 | 0.5106 |
|  | Total Cholesterol | 182.53±51.92 | 157.13±47.65 | 0.1452 |
|  | Triglyceride | 130.58±33.18 | 155.29±68.67 | 0.4072 |
| **Non-diabetic IS** | Fasting blood sugar | 102.61±12.60 | 97.1±9.65 | 0.1738 |
|  | Total Cholesterol | 173.3±51.08 | 175.08±45.08 | 0.7852 |
|  | Triglyceride | 128.36±32.84 | 152.45±47.80 | 0.1264 |
| **Non-diabetic Controls** | Fasting blood sugar | 98.29±18.59 | 87.67±19.46 | 0.3022 |
|  | Total Cholesterol | 138.08±23.63 | 137.43±36.03 | 1.000 |
|  | Triglyceride | 146.14±59.28 | 130±58.41 | 0.5388 |

### *TCF7L2* mRNA Expression in Correlation to Genotype of rs79016695

Next, the influence of rs79016695T/C on *TCF7L2* gene expression was examined among the stroke cases. By doing this, we observed significant difference in mean gene expression for its ‘T’ allele-carrying genotypes (TC+CC = -2.312 ± 0.1323, n = 8; TT = -2.821 ± 0.09848, n = 16) between the groups [Figure 3].

**Figure 3:**
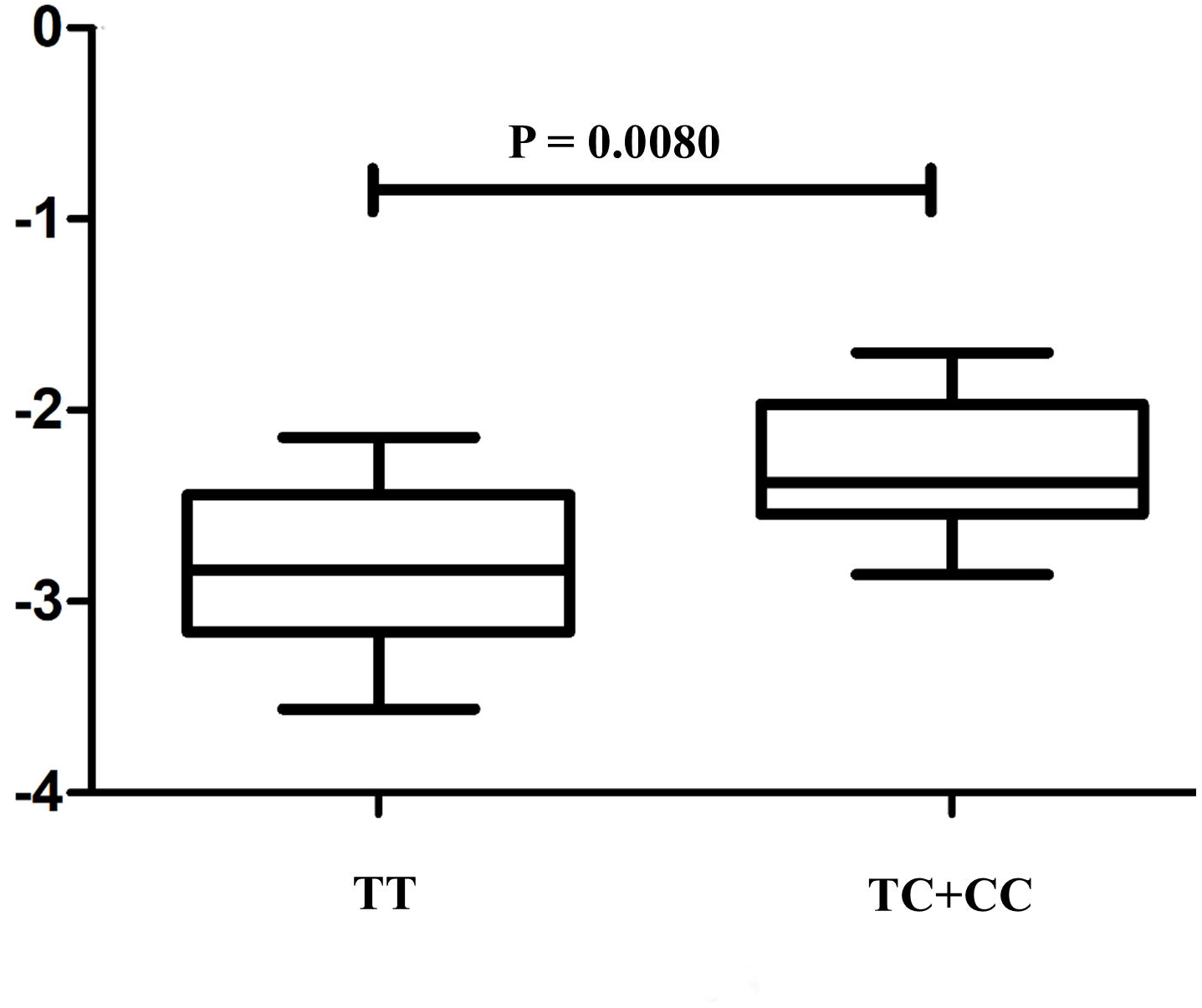
Comparison of log_2_-transformed *TCF7L2* mRNA expression levels among individuals grouped by rs7901695 T/C genotypes.

## Discussion

This study provides important insights into the role of the Transcription factor 7-like 2 (*TCF7L2*) gene in the pathogenesis of ischemic stroke, particularly in the context of metabolic disturbances such as diabetes and dyslipidemia.

A key finding is the significant downregulation of *TCF7L2* expression in peripheral blood mononuclear cells (PBMCs) of ischemic stroke patients compared to controls, irrespective of diabetic status. This suggests that dysregulation of *TCF7L2* may be a common molecular mechanism contributing to stroke risk, potentially through its involvement in glucose and lipid homeostasis. Interestingly, we observed a significant negative correlation between *TCF7L2* expression and total cholesterol levels among stroke patients, but not with fasting blood sugar. This implies that the downregulation of *TCF7L2* may be more closely linked to dyslipidemia than hyperglycemia in the context of stroke pathogenesis. This is further supported by our finding of significantly lower *TCF7L2* expression in stroke cases with hyperlipidemia compared to those without. These results are consistent with a recent study in mice, which reported that *TCF7L2* deficiency can perturb whole-body lipid metabolism (10). Regarding the genetic aspects, our analysis did not identify any pathogenic coding variants in the *TCF7L2* gene among Indian Ischemic stroke patients. However, the genetic association analysis revealed that the ‘C’ allele and ‘CC/TC’ genotype of the rs7901695 variant were associated with an increased risk of diabetes-related ischemic stroke, but not non-diabetic stroke. Interestingly, the ‘T’ allele of rs7901695 was overrepresented in stroke patients with hyperlipidemia compared to those with normal cholesterol levels. Furthermore, the ‘T’ allele-carrying genotypes of rs7901695 showed lower *TCF7L2* expression compared to the ‘CC’ genotype among stroke cases. These findings suggest a complex interplay between *TCF7L2* genetics, expression, and the metabolic disturbances that contribute to stroke risk.

An important strength of this study is the use of PBMCs as the biospecimen for *TCF7L2* expression analysis. PBMCs are known to have considerable overlap in gene expression with brain tissue and can serve as a useful proxy for investigating molecular mechanisms in stroke (11). Additionally, we were able to demonstrate the potential diagnostic value of *TCF7L2* expression for Ischemic Stroke, with an area under the receiver operating characteristic (ROC) curve of 0.8346.

In consistent with a very recent study where TCF7L2-KO mice were found to exhibit compromised β-catenin signalling and concomitant higher cholesterol biosynthesis (12), here we provide direct evidence showing alteration in *TCF7L2* leading to stroke pathogenesis is more associated with hyperlipidemia than diabetes with genetic contribution from rs7901695 among Indians. Despite such new findings, the limitation of our study includes the representation of only patients from West Bengal and Gujarat, this may not completely represent all Indians, as India is a large country with a population diversity. Also, our study is a case-control study and its findings would require validation in a standard disease model.

In conclusion, our findings indicate that the downregulation of *TCF7L2* and genetic variants in this gene may contribute to Ischemic Stroke risk, particularly in the context of metabolic disturbances such as diabetes and hyperlipidemia (Figure 4). Further studies are warranted to elucidate the precise molecular pathways by which *TCF7L2* dysregulation influences stroke pathogenesis, including validation in appropriate disease models and examination of trans-regulatory mechanisms.

**Figure 4:**
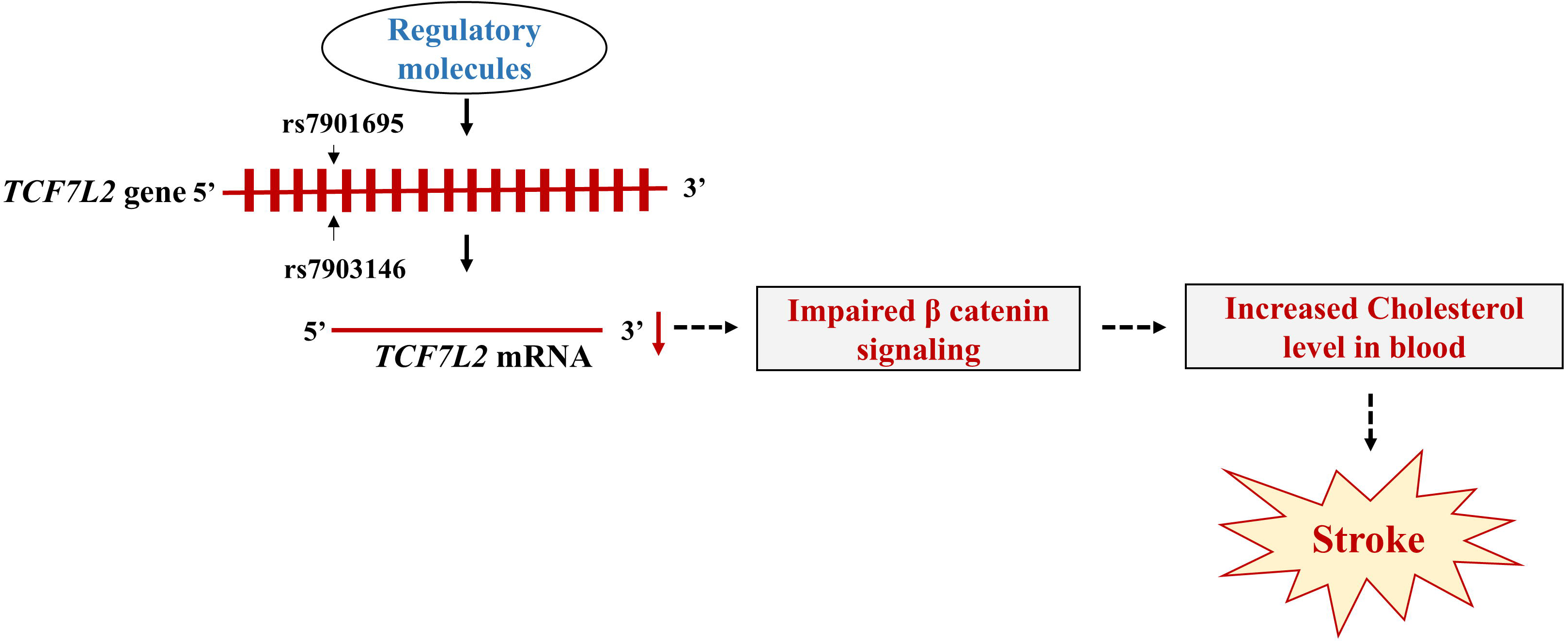
Schematic representation describing *TCF7L2* mediated stroke pathogenesis through dyslipidaemia.

## Ethics Approval

All procedures performed in studies involving human participants were in accordance with the ethical standards of the National Neurosciences Centre Calcutta, Kolkata, India, Nil Ratan Sircar Medical College & Hospital, Kolkata, India and Shree Krishna Hospital, Pramukhswami Medical College, Karamsad, Gujrat, India. The Ethics Committees of the abovementioned Institutes approved the study protocol. Informed consent was taken as per guidelines of the Indian Council of Medical Research, National Ethical Guidelines for Biomedical and Health Research involving human participants, India.

## Acknowledgement

The authors thank the patients who participated in the study.

## Authors contribution

DS and AB were responsible for study design, experimental work, data analysis, manuscript writing, and funding. AR, EB has done experimental work, data analysis. SN, DR, has done biochemical analysis and manuscript preparation. JM, KCG, TKB, PK, SD have done Clinical diagnosis, and manuscript preparation. SKC and VAP have done sample collection and clinical data management. SPH provided instrumentation facility and his valuable input for manuscript preparation. SG contributed in accumulation of funding and Manuscript preparation.

All authors read the draft, provided their inputs, and agreed on the final version of the manuscript.

## Consent to participate

All the authors participated and contributed significantly to this research work.

## Consent for publication

All the authors gave their consent to publish this research work in the Journal Archives of Medical Research

## Data Availability Statement

The data described in this study are available from the corresponding author upon reasonable request.

## Informed consent

Informed consent from all the participants was received before clinical data and sample collection.

## Statements and Declarations

The article is original and has not been published previously. All the stated authors approve its submission. The authors declare that there are no relevant financial or nonfinancial interests to disclosure.

## References

1. Mosenzon O, Cheng AY, Rabinstein AA, Sacco S. Diabetes and Stroke: What Are the Connections? J Stroke 2023;25(1):26–38. doi: 10.5853/jos.2022.02306.

2. Georgakis MK, Harshfield EL, Malik R, et al. Diabetes Mellitus, Glycemic Traits, and Cerebrovascular Disease: A Mendelian Randomization Study. Neurology 2021;96(13):e1732–e1742. doi: 10.1212/WNL.0000000000011555.

3. Del Guerra S, Lupi R, Marselli L, et al. Functional and molecular defects of pancreatic islets in human type 2 diabetes. Diabetes 2005;54(3):727–735. doi:10.2337/diabetes.54.3.727

4. Grant SF, Thorleifsson G, Reynisdottir I, et al. Variant of transcription factor 7-like 2 (TCF7L2) gene confers risk of type 2 diabetes. Nat Genet. 2006;38(3):320–323.

5. Choi HJ, Lee DH, Jeon HJ, Kim DS, Lee YH, Oh T. Transcription factor 7-like 2 (TCF7L2) gene polymorphism rs7903146 is associated with stroke in type 2 diabetes patients with long disease duration. Diabetes Res Clin Pract 2014;103(3):e3–6. doi: 10.1016/j.diabres.2013.12.051.

6. Verma AK, Ali Beg MM, Saleem M, et al. Cell-free TCF7L2 gene alteration and their association with Type 2 diabetes mellitus in North Indian population. Meta Gene 2020;25:100727.doi10.1016/j.mgene.2020.100727.

7. Gunavathy N, Balaji R, Kumaravel V. Association of TCF7L2 Variants in Type 2 Diabetes Mellitus with Hypertriglyceridemia - A Case-Control Study. Indian J Endocrinol Metab 2023;27(4):346–350. doi: 10.4103/ijem.ijem_35_23.

8. Geoghegan G, Simcox J, Seldin MM, et al. Targeted deletion of Tcf7l2 in adipocytes promotes adipocyte hypertrophy and impaired glucose metabolism. Mol Metab 2019;24:44–63. doi: 10.1016/j.molmet.2019.03.003

9. Roche Diagnostics. Cobas c-501 Analyzer: Technical Manual. Basel, Switzerland: Roche Diagnostics; 2020.

10. Nguyen-Tu MS, Martinez-Sanchez A, Leclerc I, Rutter GA, da Silva Xavier G. Adipocyte-specific deletion of Tcf7l2 induces dysregulated lipid metabolism and impairs glucose tolerance in mice. Diabetologia 2021;64(1):129–141. doi:10.1007/s00125-020-05292-4

11. Moore DF, Altarescu G, Barker WC, Patronas NJ, Herscovitch P, Schiffmann R. Using peripheral blood mononuclear cells to measure nuclear factor-kappa B activity in ischemic stroke patients. Cell Mol Neurobiol. 2005;25(6):1105–1109.

12. Szewczyk LM, Lipiec MA, Liszewska E, et al. Astrocytic β-catenin signaling via TCF7L2 regulates synapse development and social behavior. Mol Psychiatry 2024;29(1):57–73. doi:10.1038/s41380-023-02281-y

